# The oxytocin system in patients with craniopharyngioma: A systematic review

**DOI:** 10.1101/2024.07.31.24311260

**Authors:** Amy Mann, Jennifer Kalitsi, Khushali Jani, Daniel Martins, Ritika R Kapoor, Yannis Paloyelis

**Author notes:** **Correspondence:** Amy Mann, Department of Neuroimaging (P089), Institute of Psychiatry, Psychology and Neuroscience, De Crespigny Park, SE5 8AF, King’s College London, UK. These authors share senior authorship (Ritika R Kapoor, Yannis Paloyelis).

## Abstract

Craniopharyngioma is a benign tumour affecting the hypothalamic and pituitary regions, which are involved in the production and secretion of oxytocin. We conducted a systematic review to assess dysregulation of the oxytocin system in craniopharyngioma and associations with neurobehavioural, eating, and metabolic abnormalities. Eight studies (n=72 patients) were included. Evidence for dysfunction of the endogenous oxytocin system in craniopharyngioma is limited and mixed. While no significant differences in baseline salivary oxytocin concentrations were reported between patients with craniopharyngioma and controls, patients with craniopharyngioma were found to have blunted salivary oxytocin response following exercise stimulation and this was associated with greater state anxiety and higher BMI. Studies administering exogenous oxytocin are sparse and do not meet required standards. Hypothalamic damage may pose an additional mechanism of oxytocin dysregulation. Improving understanding of the oxytocin system in craniopharyngioma could be pivotal for exploring the potential therapeutic role of exogenous oxytocin in this condition.

## 1 Introduction

Craniopharyngioma is a rare benign tumour affecting the hypothalamus and pituitary gland, with an incidence of 0.5 to 2 cases per million people per year (Nielsen et al., 2011). Peak onset is between 5-14 years old in children and 50-74 years in adults (Bunin et al., 1998), where 30-50% of all cases present during childhood or adolescence (Nielsen et al., 2011). Although craniopharyngioma is a histologically benign tumour, patients experience significant morbidity related to local infiltration of surrounding structures by the tumour and because of the treatment strategies, which involve resection of the tumour and/ or radiotherapy (Müller, 2010). The long-term morbidities impairing quality of life of these patients include varying degrees of hypopituitarism, and visual and neurological deficits (Müller, 2020; Zhou et al., 2021). Cognitive-behavioural, and emotional difficulties (hitherto referred to as neurobehavioural impairment) (Özyurt et al., 2015; Zada et al., 2013), hyperphagia (i.e., pathological overeating), and obesity (Roth, 2011) are additional prevalent manifestations in these patients. Impairments may persist following treatment of the tumour (Mende et al., 2020), and often increase in severity, likely as a consequence of post-operative hypothalamic damage. At present, there is no standard of care for neurobehavioural impairment or hyperphagic eating behaviours experienced by affected patients with craniopharyngioma, despite posing a significant challenge for both patients and their families.

Despite correcting other hormone deficiencies, disruption of the oxytocin system and the potential benefits of the administration of exogenous oxytocin are yet to be assessed, and thus not considered in routine care for patients with craniopharyngioma. Oxytocin is a hypothalamic neuropeptide primarily synthesised in the magnocellular and parvocellular neurons of the paraventricular and supraoptic nuclei of the hypothalamus. Magnocellular neurons project to the posterior pituitary for oxytocin release into peripheral circulation, whilst both magnocellular and parvocellular neurons are involved in central oxytocin release (Althammer & Grinevich, 2017). Oxytocin is known to be implicated in multiple physiological and behavioural pathways including the regulation of social-cognitive functioning (Johnson & Young, 2017), the modulation of feeding behaviour (Lawson, 2017), and neuroinflammation (Knoop et al., 2022). As such, oxytocin dysregulation has been suggested in a number of neurodevelopmental and psychiatric conditions, including autism spectrum disorder (ASD) (John & Jaeggi, 2021), schizophrenia, and anorexia nervosa (Ferreira & Osório, 2022). In addition, the anorexigenic effects of oxytocin, with reductions in food intake, weight and fat, and improvements in glucose homeostasis, have been observed in pre-clinical (e.g., Blevins et al. (2015); see also Leslie et al. (2018) and clinical studies (Lawson, 2017). Interest in exogenous oxytocin as a therapeutic for this group is therefore motivated by the potential to benefit patients with craniopharyngioma across key neurobehavioural and metabolic clinical features.

Damage to the hypothalamo-pituitary region, which is a common feature of craniopharyngioma, poses a likely mechanism of disruption of the homeostatic regulation of physiological concentrations of oxytocin centrally and peripherally, and/or its central and peripheral release in response to stimulation. Therefore, hypothalamo-pituitary damage may have considerable implications for metabolic and neurobehavioural functioning in craniopharyngioma. Specifically, there are different degrees of hypothalamic damage, caused pre-operatively and/or post-operatively, involving the anterior hypothalamic regions (grade I), and extending to the posterior hypothalamic regions with or without mammillary body involvement (grade II) (Müller et al., 2012; Müller et al., 2019). Studies have shown that the cystic and solid components of craniopharyngioma are high in lipids, cholesterol, and pro-inflammatory markers (Apps et al., 2018; Whelan et al., 2020), where the cystic fluid has shown to initiate an inflammatory activation of the microglia, causing damage to the hypothalamus (Ainiwan et al., 2022). This lipid-rich and inflammatory composition is greater than that seen in other benign tumours (Apps et al., 2018; Donson et al., 2017), likely accounting for the prevalent hypothalamic dysfunction in this specific tumour type (alongside the damage caused by surgical excision and/or radiotherapy). The degree of hypothalamic involvement predicts outcome type and severity in craniopharyngioma (Müller, 2020). The presence of hypothalamic damage (caused pre-operatively and/or post-operatively) therefore poses a key clinical feature in need of consideration in patients with craniopharyngioma as it contributes to clinical heterogeneity and heterogeneity in the degree of involvement of the oxytocin system.

This is the first systematic review that aims to assess the extent to which the oxytocin system is compromised in craniopharyngioma, the relevance of hypothalamic damage, and whether alterations in the function of the oxytocin system may be associated with the neurobehavioural and metabolic dysfunction observed in this condition. It is anticipated that improving understanding of the involvement of the oxytocin system in craniopharyngioma could be pivotal for exploring the potential therapeutic role of exogenous oxytocin in this condition.

## 2 Methods

The present systematic review was pre-registered with PROSPERO (ID: CRD42023397966) and followed the Preferred Reporting Items for Systematic Reviews and Meta-Analyses (PRISMA) guidelines (Moher et al., 2009; Page et al., 2021) (see Supplementary Material Table S1).

### 2.1 Search strategy

PubMed, Embase, and PsycInfo were searched to identify peer-reviewed articles published in English, from inception through January 19, 2024. The following search terms were used and adjusted based on the requirements of each database: (oxytocin OR OT OR OXT OR OXTR OR CD38) AND (craniopharyngioma). The Cochrane Central Register of Controlled Trials was also searched using the terms “craniopharyngioma” and “oxytocin”. No filters or limits were applied to the search. Hand-searching of the reference lists of included studies and relevant literature reviews was performed to search for additional studies.

### 2.2 Study selection

Articles were exported into Rayyan (http://rayyan.qcri.org) where duplicate articles were removed using the duplicate identification tool. Authors AM and JK/ KJ independently reviewed the titles and abstracts against the inclusion and exclusion criteria. Due to the novelty of this field, our inclusion criteria were intentionally broad and included original peer-reviewed articles with: 1) a sample of humans with craniopharyngioma; and 2) assessment of the oxytocin system (e.g., baseline, pre- and/ or post-intervention for release of endogenous oxytocin, pre- and/ or post-exogenous oxytocin intervention, or genetic association); and 3) measurement of neurobehavioural outcomes (e.g., behavioural, cognitive, social, emotional, psychiatric) or eating behaviours (e.g., hyperphagia); or 4) measurement of metabolic outcomes (e.g., body mass index; BMI). Clinical trial registrations were included and extracted where sufficient outcome data had been reported. Non-English articles and those where full-texts could not be obtained were excluded due to the inability to extract required data. All excluded articles were documented in an Excel database with justifications for exclusion.

### 2.3 Data charting and synthesis

Data was independently extracted by three authors, AM, JK and KJ, into a data extraction spreadsheet generated during protocol development. The following data were extracted from included papers: 1) first author, 2) year of publication, 3) study design and sample size, 4) age and gender of participants, 5) participant clinical characteristics (e.g., endocrine morbidity, visual impairment, grade of hypothalamic damage), 6) treatment status (i.e., if pre- or post-operative), 7) comparator group demographics (if applicable), 8) information on measurement, sampling, and quantification of oxytocin, or oxytocin treatment 9) information on neurobehavioural or metabolic outcomes, or eating behaviours 10) associations between the oxytocin system and neurobehavioural or metabolic outcomes, or eating behaviours, and 11) group differences between craniopharyngioma and comparators in the oxytocin system and neurobehavioural or metabolic outcomes, or eating behaviours. Information on demographic, clinical, and outcome data are detailed in Table 1. A narrative synthesis was used to integrate the key findings of the included articles.

**Table 1.**
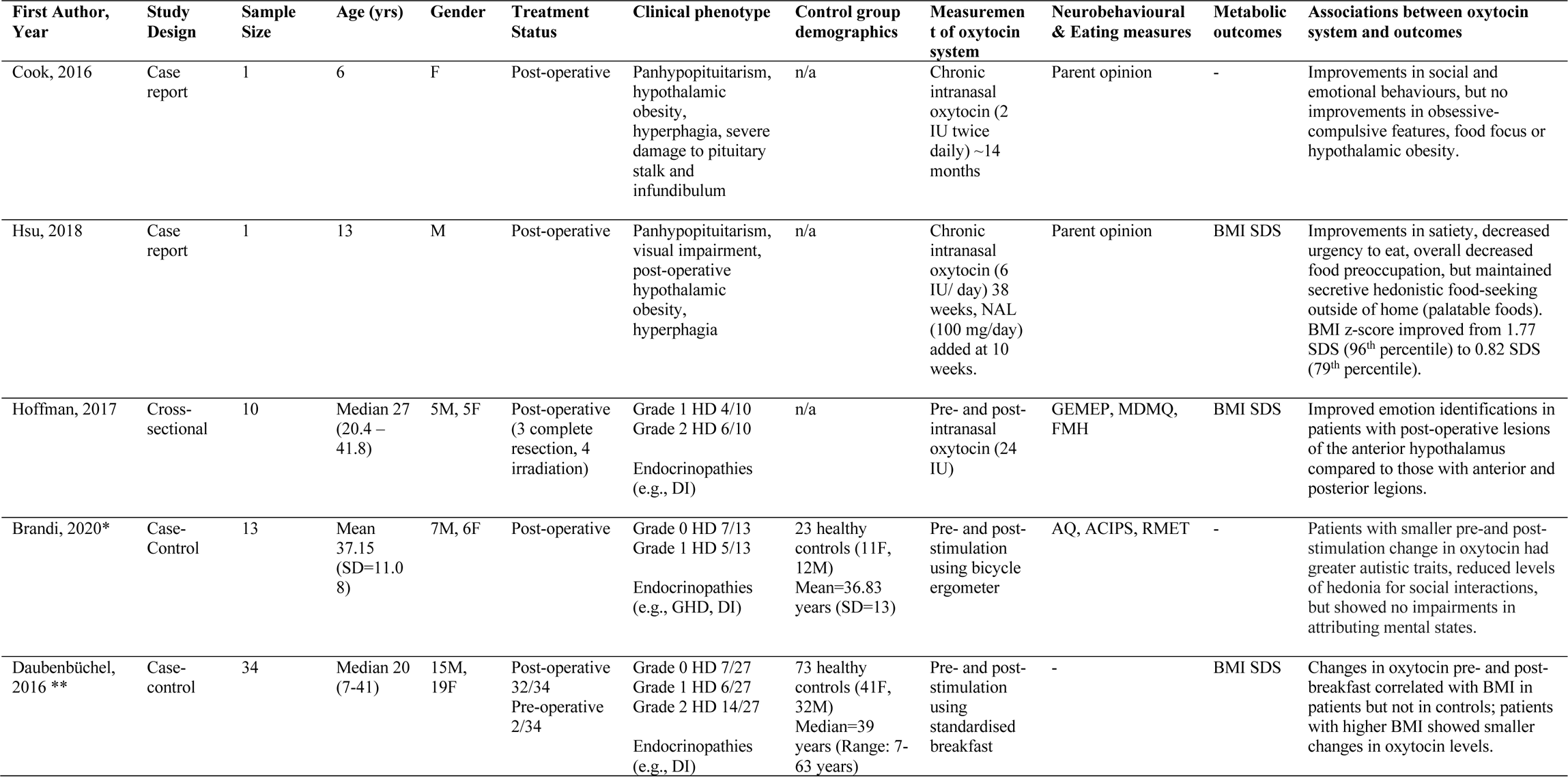
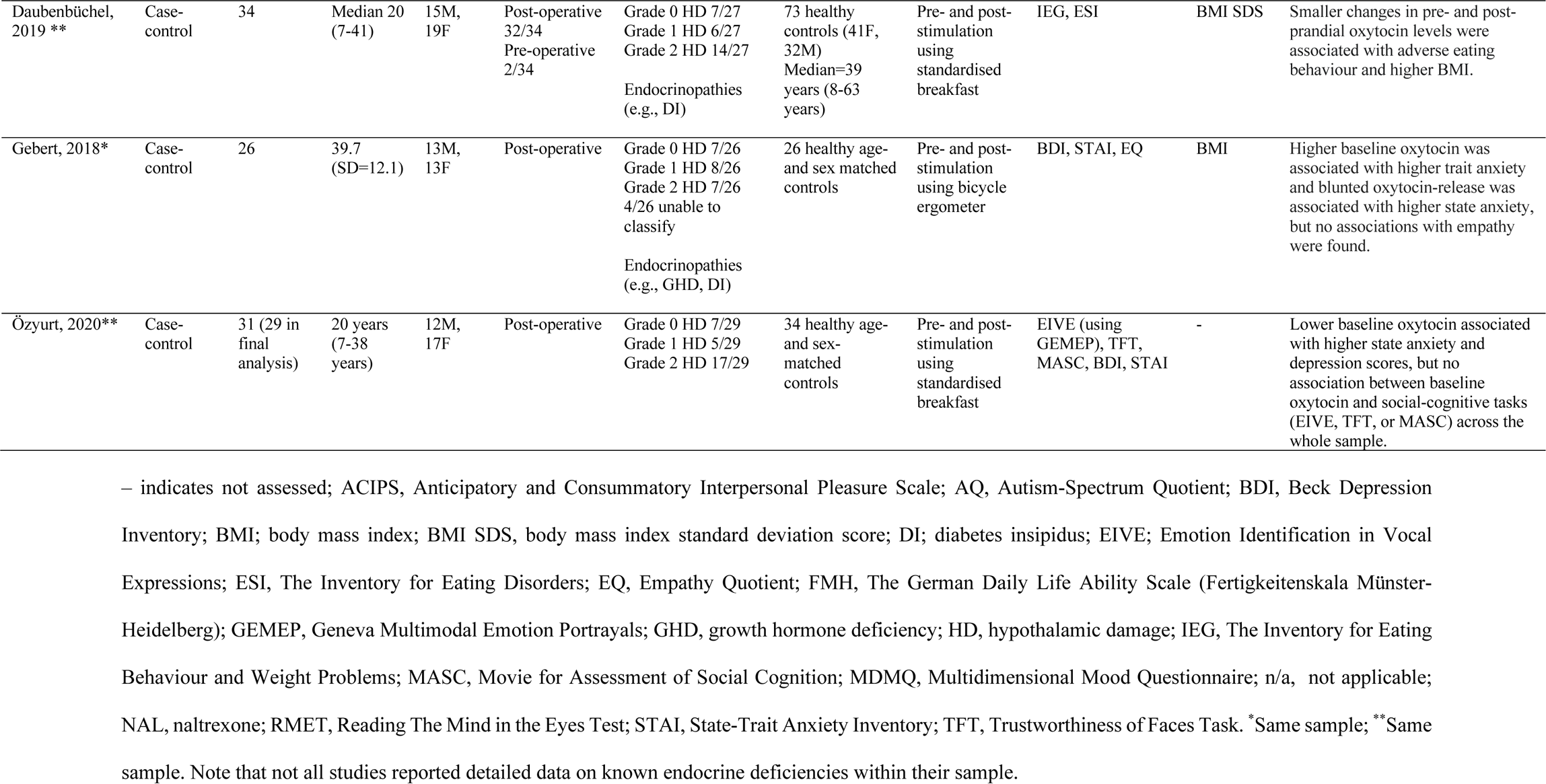
Clinical, demographic, and key outcome data for included studies.

| First Author, Year | Study Design | Sample Size | Age (yrs) | Gender | Treatment Status | Clinical phenotype | Control group demographics | Measurement of oxytocin system | Neurobehavioural & Eating measures | Metabolic outcomes | Associations between oxytocin system and outcomes |
| --- | --- | --- | --- | --- | --- | --- | --- | --- | --- | --- | --- |
| Cook, 2016 | Case report | 1 | 6 | F | Post-operative | Panhypopituitarism, hypothalamic obesity, hyperphagia, severe damage to pituitary stalk and infundibulum | n/a | Chronic intranasal oxytocin (2 IU twice daily) ~14 months | Parent opinion | - | Improvements in social and emotional behaviours, but no improvements in obsessive-compulsive features, food focus or hypothalamic obesity. |
| Hsu, 2018 | Case report | 1 | 13 | M | Post-operative | Panhypopituitarism, visual impairment, post-operative hypothalamic obesity, hyperphagia | n/a | Chronic intranasal oxytocin (6 IU/ day) 38 weeks, NAL (100 mg/day) added at 10 weeks. | Parent opinion | BMI SDS | Improvements in satiety, decreased urgency to eat, overall decreased food preoccupation, but maintained secretive hedonistic food-seeking outside of home (palatable foods). BMI z-score improved from 1.77 SDS (96<sup>th</sup> percentile) to 0.82 SDS (79<sup>th</sup> percentile). |
| Hoffman, 2017 | Cross-sectional | 10 | Median 27 (20.4 – 41.8) | 5M, 5F | Post-operative (3 complete resection, 4 irradiation) | Grade 1 HD 4/10<br>Grade 2 HD 6/10<br>Endocrinopathies (e.g., DI) | n/a | Pre- and post-intranasal oxytocin (24 IU) | GEMEP, MDMQ, FMH | BMI SDS | Improved emotion identifications in patients with post-operative lesions of the anterior hypothalamus compared to those with anterior and posterior lesions. |
| Brandi, 2020* | Case-Control | 13 | Mean 37.15 (SD=11.08) | 7M, 6F | Post-operative | Grade 0 HD 7/13<br>Grade 1 HD 5/13<br>Endocrinopathies (e.g., GHD, DI) | 23 healthy controls (11F, 12M)<br>Mean=36.83 years (SD=13) | Pre- and post-stimulation using bicycle ergometer | AQ, ACIPS, RMET | - | Patients with smaller pre-and post-stimulation change in oxytocin had greater autistic traits, reduced levels of hedonia for social interactions, but showed no impairments in attributing mental states. |
| Daubenbüchel, 2016 ** | Case-control | 34 | Median 20 (7-41) | 15M, 19F | Post-operative 32/34<br>Pre-operative 2/34 | Grade 0 HD 7/27<br>Grade 1 HD 6/27<br>Grade 2 HD 14/27<br>Endocrinopathies (e.g., DI) | 73 healthy controls (41F, 32M)<br>Median=39 years (Range: 7-63 years) | Pre- and post-stimulation using standardised breakfast | - | BMI SDS | Changes in oxytocin pre- and post-breakfast correlated with BMI in patients but not in controls; patients with higher BMI showed smaller changes in oxytocin levels. |
| Daubenbüchel, 2019 ** | Case-control | 34 | Median 20 (7-41) | 15M, 19F | Post-operative 32/34<br>Pre-operative 2/34 | Grade 0 HD 7/27<br>Grade 1 HD 6/27<br>Grade 2 HD 14/27<br>Endocrinopathies (e.g., DI) | 73 healthy controls (41F, 32M)<br>Median=39 years (8-63 years) | Pre- and post-stimulation using standardised breakfast | IEG, ESI | BMI SDS | Smaller changes in pre- and post-prandial oxytocin levels were associated with adverse eating behaviour and higher BMI. |
| Gebert, 2018* | Case-control | 26 | 39.7 (SD=12.1) | 13M, 13F | Post-operative | Grade 0 HD 7/26<br>Grade 1 HD 8/26<br>Grade 2 HD 7/26<br>4/26 unable to classify<br>Endocrinopathies (e.g., GHD, DI) | 26 healthy age- and sex matched controls | Pre- and post-stimulation using bicycle ergometer | BDI, STAI, EQ | BMI | Higher baseline oxytocin was associated with higher trait anxiety and blunted oxytocin-release was associated with higher state anxiety, but no associations with empathy were found. |
| Özyurt, 2020** | Case-control | 31 (29 in final analysis) | 20 years (7-38 years) | 12M, 17F | Post-operative | Grade 0 HD 7/29<br>Grade 1 HD 5/29<br>Grade 2 HD 17/29 | 34 healthy age- and sex-matched controls | Pre- and post-stimulation using standardised breakfast | EIVE (using GEMEP), TFT, MASC, BDI, STAI | - | Lower baseline oxytocin associated with higher state anxiety and depression scores, but no association between baseline oxytocin and social-cognitive tasks (EIVE, TFT, or MASC) across the whole sample. |

– indicates not assessed; ACIPS, Anticipatory and Consummatory Interpersonal Pleasure Scale; AQ, Autism-Spectrum Quotient; BDI, Beck Depression Inventory; BMI; body mass index; BMI SDS, body mass index standard deviation score; DI; diabetes insipidus; EIVE; Emotion Identification in Vocal Expressions; ESI, The Inventory for Eating Disorders; EQ, Empathy Quotient; FMH, The German Daily Life Ability Scale (Fertigkeitskala Münster-Heidelberg); GEMEP, Geneva Multimodal Emotion Portrayals; GHD, growth hormone deficiency; HD, hypothalamic damage; IEG, The Inventory for Eating Behaviour and Weight Problems; MASC, Movie for Assessment of Social Cognition; MDMQ, Multidimensional Mood Questionnaire; n/a, not applicable; NAL, naltrexone; RMET, Reading The Mind in the Eyes Test; STAI, State-Trait Anxiety Inventory; TFT, Trustworthiness of Faces Task. \*Same sample; \*\*Same sample. Note that not all studies reported detailed data on known endocrine deficiencies within their sample.

**Table 2.** Methods of assessing the endogenous oxytocin system in craniopharyngioma and healthy controls.

| First Author, Year | Oxytocin Measurement | Sampling | Sample preparation | Sample Extraction | Oxytocin Quantification | Basal OT-levels in pg/mL | Post-intervention OT-levels in pg/mL | ΔOSC (pre- post), pg/mL |
| --- | --- | --- | --- | --- | --- | --- | --- | --- |
| Hoffman, 2017 | Basal:<br>Post- single intranasal administration of 24 IU oxytocin (Syntocinon® Spray, Novartis, Basel, Switzerland): 3 puffs per nostril.<br>Saliva: 40 min after<br>Urine 90 mins after | Saliva, urine | NR | NR | RIA (RIAGnosis, Sinzing, Germany) | Median (Range)<br>Saliva = 0.32 (0.25–3.60)<br>Urine = 0.90 (0.42–1.59) | *Post-Intranasal Oxytocin:*<br>Median (Range)<br>Saliva = 87.3 (5.21 – 97.27)<br>Urine = 11.13 (1.32-105.68) | - |
| Brandi, 2020* | Basal: AM (8:30am start, fasting state, food >12 h, water >1 h)<br>Post-stimulation: bicycle ergometer | Saliva | Centrifuged at 3000g for 5min at 4°C, then stored at -20°C. | Yes – all samples extracted and assayed in same batch at same time to eliminate inter-assay variation. | RIA (RIAGnosis, Sinzing, Germany) | Mean (SD)<br>CP = 1.90 (1.43)<br>HC = 1.24 (1.08)<br>*p*=0.865 | NR | Mean (SD)<br>CP = -7.90% (20.6)<br>HC = 21.26% (27.41)<br>*p*<.001 |
| Daubenbüchel, 2016 ** | Basal: AM<br>Post-stimulation: 60 mins after standardised breakfast (approx. 10–15 kcal/kg body weight; 8am) | Saliva | Placed immediately on ice, centrifuged, then stored frozen. A protease inhibitor was not used in centrifugation. | Yes – all samples extracted. | EIA | Median (Range)<br>CP = 3.61 (0.07 – 12.45)<br>HC = 3.35 (0.06 – 15.33)<br>*n.s.* | Median (Range)<br>CP = 3.18 (0.07–11.24)<br>HC = 2.78 (0.06–22.68)<br>*n.s.* | Median (Range)<br>CP = -0.93 (-6.03-8.2)<br>HC = -0.34 (-11.6-13.99)<br>*n.s.* |
| Daubenbüchel, 2019 ** | Basal: AM<br>Post-stimulation: 60 mins after standardised breakfast (approx. 10–15 kcal/kg body weight; 8am) | Saliva | Placed immediately on ice, centrifuged, then stored frozen. A protease inhibitor was not used in centrifugation. | Yes – all samples extracted. | EIA | Median (Range)<br>CP = 3.6 (0.1-12.5)<br>HC = 3.4 (0.1-15.3) | Median (Range)<br>CP = 3.2 (0.1-11.2)<br>HC = 2.8 (0.1-22.7) | Median (Range)<br>CP = -0.9 (-6.0-8.2)<br>HC = -0.3 (-11.6-14.0) |
| Gebert, 2018* | Basal: AM (8:30am start, fasting state, food >12 h, water >1 h)<br>Post-stimulation: bicycle ergometer | Saliva | Centrifuged at 3000g for 5min at 4°C, then stored at -20°C. | Yes – all samples extracted and assayed in same batch at same time to eliminate inter-assay variation. | RIA (RIAgnosis, Sinzing, Germany) | Mean (SD)<br>CP = 1.46 (1.20)<br>HC = 1.33 (1.13)<br>*p*=0.731 | Mean (SD)<br>CP = 1.26 (0.87)<br>HC = 1.66 (1.76)<br>*p*= 0.391 | Mean<br>CP = -13.7%<br>HC = 24.8%<br>(GLM time x group)<br>*p*=0.005 |
| Özyurt, 2020** | Basal | Saliva | Placed immediately on ice, centrifuged, then stored frozen until analysis. A protease inhibitor was not used in centrifugation. | Yes – all samples extracted. | EIA | Median (IQR)<br>CP = 3.3 (3.6)<br>HC = 4.42 (7.6)<br>*p*=0.329 | - | - |

– indicates, not assessed; EIA, enzyme immunoassay; CP, craniopharyngioma; HC, healthy control; IQR, interquartile range; NR, not reported; n.s., not significant (no *p*-value reported by authors); RIA, radioimmunoassay; SD, standard deviation. \*Same sample; \*\*Same sample.

### 2.4 Quality assessment

Due to the variance in study designs, appropriate versions of the JBI Critical Appraisal checklists were used to assess individual study quality (see Supplementary Material Figure S1 to 3). Adjustments to the JBI checklists were made by the study team for studies administering exogenous oxytocin. Two independent reviewers assessed the quality of each study; any discrepancies were resolved through discussion or intervention by a third reviewer. No study was excluded due to a poor-quality assessment.

## 3 Results

The study selection process is detailed in Figure 1. The search yielded 67 unique articles, of which eight studies were included. Of the eight included studies, data of 72 patients are reported on across two case reports (Cook et al., 2016; Hsu et al., 2017), one interventional study administering a single dose of 24IU intranasal oxytocin (Hoffmann et al., 2017), and five cross-sectional, case-control studies (Brandi et al., 2020; Daubenbüchel et al., 2016; Daubenbüchel et al., 2019; Gebert et al., 2018; Özyurt et al., 2020). No papers assessing genetic associations were found from the search.

**Figure 1.**
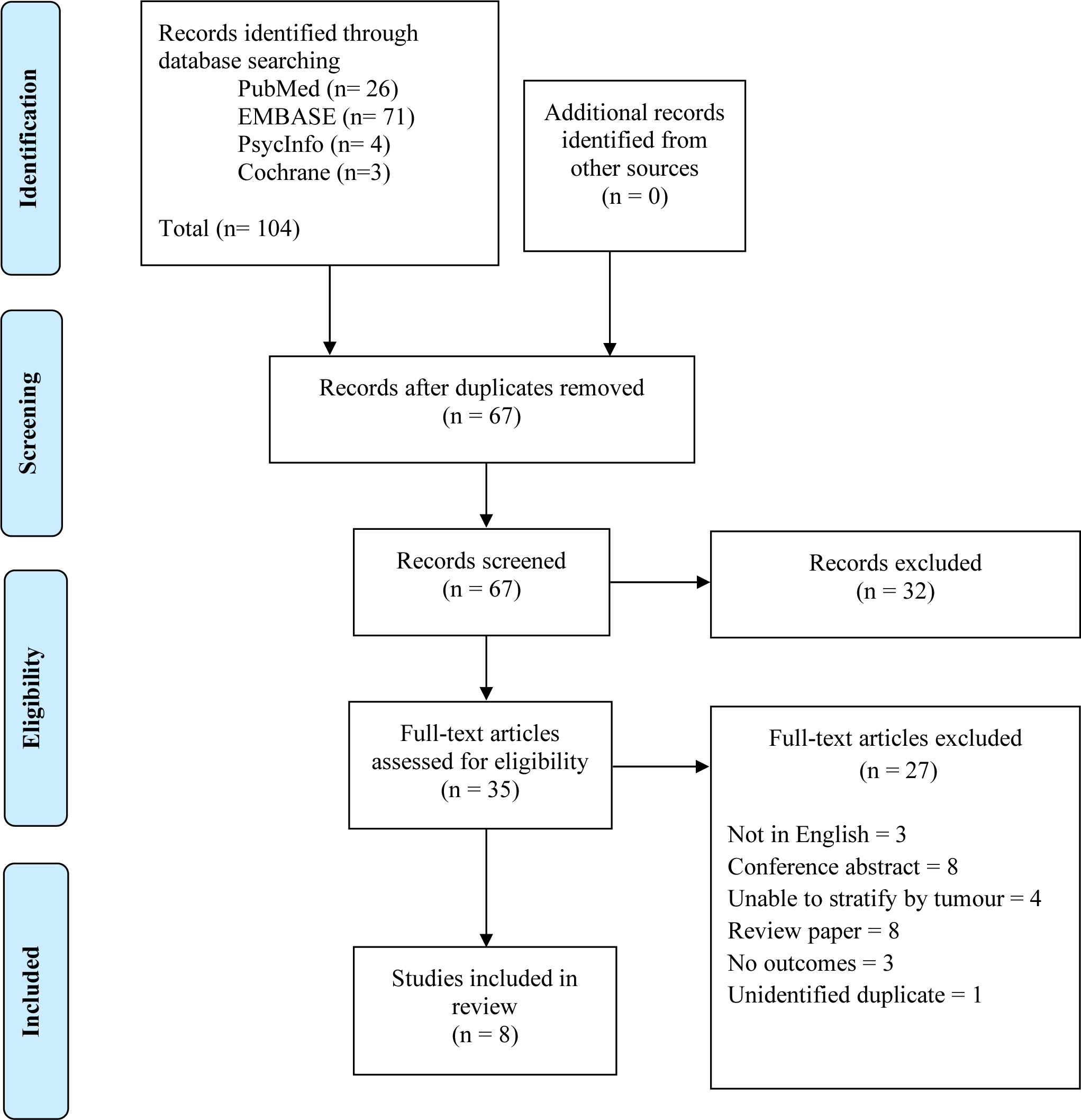
PRISMA diagram of search strategy

It is important to note that oxytocin samples were collected from the same participant sample for Daubenbüchel et al. (2016) and Daubenbüchel et al. (2019); Özyurt et al. (2020) reports on a subsample of these studies. Similarly, Brandi et al. (2020) reports oxytocin data on a subsample of Gebert et al. (2018). Therefore, only unique results are discussed in this review and the earliest study will be referenced for studies reporting on the same data.

### 3.1 Methods of assessing the oxytocin system

Varied approaches to assessing the oxytocin system in craniopharyngioma were adopted in the eight included studies (see Table 1). Two case reports assessed the effects of long-term use of low dose (4IU-6IU/ day) intranasal oxytocin on parent-reported behavioural change and BMI (Cook et al., 2016; Hsu et al., 2017). One study assessed the effects of a single-dose of intranasal oxytocin on emotion recognition performance (Hoffmann et al., 2017). Two research groups assessed concentrations of endogenous oxytocin before (baseline) and after an intervention intended to activate the endogenous oxytocin system, in comparison to a healthy control group (Daubenbüchel et al., 2016; Gebert et al., 2018). Özyurt et al. (2020) reported only on baseline measurements of fasting salivary oxytocin in comparison to healthy controls. All studies assessing endogenous oxytocin concentrations measured salivary oxytocin (Daubenbüchel et al., 2016; Gebert et al., 2018; Hoffmann et al., 2017), with one study also measuring oxytocin in urine (Hoffmann et al., 2017). Two research groups quantified oxytocin using radioimmunoassay (RIA) (Gebert et al., 2018; Hoffmann et al., 2017), and one group used enzyme immunoassay (EIA) (Daubenbüchel et al., 2016).

Across the two research groups (Daubenbüchel et al., 2016; Gebert et al., 2018) implementing a pre- and post-activation paradigm, baseline oxytocin concentrations were obtained in fasting state between 08:00 – 08:30 as a single salivary sample (using Salivettes). In terms of activating the oxytocin system, Gebert et al. (2018) used a bicycle ergometer where stepwise increasing wattage difficulty was used and participants continued until exertion (lactate in capillary blood was measured repeatedly to standardise for individual exertion). Participants exercised on the bike for up to 7 minutes and stopped when lactate levels >4 mmol/l or if lactate levels were maintained at 4 mmol/l, when participants reached physical exhaustion, or after 25 minutes of exercise. A single salivary sample was taken to measure oxytocin immediately after exhaustion was reached. By contrast, Daubenbüchel et al. (2016) activated the endogenous oxytocin system by administering a standardised breakfast meal (10 to 15 kcal/kg body weight) and post-prandial concentrations of oxytocin were measured by a single salivary sample 60 minutes after consumption of the meal.

Hoffmann et al. (2017) was the only study to implement an interventional pre- and post-intranasal oxytocin paradigm to assess the emotion identification ability in 10 adults before and at ∼60 minutes after intranasal oxytocin administration. No placebo arm or blinding was implemented in this study; all participants received a single dose of 24IU intranasal oxytocin administered using a nasal spray. The authors additionally measured baseline and post-intranasal concentrations of salivary (at 45 to 60 after administration) and urinary (at 90 minutes after administration) oxytocin.

#### 3.1.1 Intranasal oxytocin studies

Across three studies administering intranasal oxytocin, consistent improvements were found for socio-emotional functioning, with unclear evidence for metabolic benefits. One case report of a 6-year-old female found no improvements in food-related obsessive-compulsive features or weight, but did report parent-observed improvements in social and emotional behaviours over approximately 14 months (dosage of 4IU/day) (Cook et al., 2016). Another case report administering 6IU/day intranasal oxytocin in a 13-year-old male did report improvements in overall food preoccupation and BMI z-score, which decreased from 1.77 SDS (96^th^ percentile) to 0.82 SDS (79^th^ percentile) over 48 weeks (Hsu et al., 2017). The difference in effects of oxytocin on BMI/ weight here may be accounted for by the addition of naltrexone (100mg/day) over the treatment period in Hsu et al. (2017). Naltrexone is an opioid antagonist with selective preference for μ-opioid receptor binding that has shown to be an effective treatment for adult obesity (Kulak-Bejda et al., 2021); μ-opioid receptor antagonism has shown to potentiate the effects of oxytocin (Nisbett et al., 2024) and increase plasma oxytocin concentrations by disinhibition of central oxytocin release in rodents during late pregnancy (Douglas et al., 1993). However, Hsu et al. (2017) did report a decrease in BMI z-score from 1.77 SDS to 1.49 SDS (93^rd^ percentile) over the 10 weeks before naltrexone was added. This suggests that, at least in part, the improvements in BMI cannot be explained by naltrexone alone. Nevertheless, in both cases, no placebo arm was implemented and the neurobehavioural and eating behaviour observations were based on parent opinions, as opposed to measurements using validated scales.

The only study investigating the effects of single-dose intranasal oxytocin on social cognition observed a numerical improvement in emotion identification (Hoffmann et al., 2017). It reported increased percentage of correct assignment of emotional vocal expressions for patients with post-operative grade I hypothalamic damage (anterior lesions; n=4) post-treatment, compared to baseline. By contrast, minimal changes or worsened scores were reported in the patients with grade II hypothalamic damage (mammillary bodies, anterior and posterior lesions; n=6) (Hoffmann et al., 2017). The authors suggested that supplementation of oxytocin may therefore only be beneficial for patients with lesions limited to the anterior hypothalamus (Hoffmann et al., 2017). Yet, given the small sample size of this study (n=4 to 6 per group), absence of inferential statistical analysis, and the lack of placebo-controlled condition, no firm conclusions can be drawn on the differential socio-cognitive effects of intranasal oxytocin dependent on grade of hypothalamic damage.

#### 3.1.2 The endogenous oxytocin system

Evidence for dysfunction of the endogenous oxytocin system in craniopharyngioma is limited and mixed, with differential findings depending on the paradigms used to measure oxytocin concentrations (e.g., single baseline measurements vs. response to intervention) and the stratification of analyses by hypothalamic damage.

Across two research groups comparing patients with craniopharyngioma and healthy controls, no significant differences in baseline salivary oxytocin concentrations were observed (Brandi et al., 2020; Daubenbüchel et al., 2016; Daubenbüchel et al., 2019; Özyurt et al., 2020). In patients with craniopharyngioma (n=70), mean/median baseline salivary oxytocin concentrations ranged between 0.32 pg/mL and 1.90 pg/mL for samples quantified using RIA (Brandi et al., 2020; Gebert et al., 2018; Hoffmann et al., 2017), and between 3.3 pg/mL and 3.6 pg/mL for samples quantified using EIA (Daubenbüchel et al., 2016; Özyurt et al., 2020). In healthy controls (n=99), mean/median baseline salivary oxytocin concentrations ranged between 1.24 pg/mL and 1.33 pg/mL for samples quantified using RIA (Brandi et al., 2020; Gebert et al., 2018), and between 3.4 pg/mL and 4.4 pg/mL for samples quantified using EIA (Daubenbüchel et al., 2016; Özyurt et al., 2020). These values are comparable to the mean oxytocin concentrations of extracted salivary samples (adjusted for quantification assay) reported in the literature for healthy adults (e.g., see Engel et al. (2019)).

When stratifying analyses by hypothalamic damage, mixed findings for differences in baseline salivary oxytocin were observed. Specifically, there was no difference when comparing patients with grade I (n=8) hypothalamic damage and no hypothalamic damage; while patients with grade II hypothalamic damage (n=7) were found to have significantly lower baseline salivary oxytocin concentrations than patients with no hypothalamic damage (n=7) (Gebert et al., 2018). By contrast, Daubenbüchel et al. (2016) found that patients with grade I (n=6) hypothalamic damage had significantly lower baseline salivary oxytocin concentrations than patients with no hypothalamic damage (n=7); yet there was no difference between patients with grade II (n=14) hypothalamic damage and no hypothalamic damage. In the same study (Daubenbüchel et al., 2016), patients with grade I hypothalamic damage were found to have lower baseline salivary oxytocin concentrations than grade II patients. A trend towards lower baseline urinary oxytocin concentrations in patients with grade I (n=4) damage compared to grade II (n=6) hypothalamic damage (*p*=0.06) was similarly reported in Hoffman et al. (Hoffmann et al., 2017). Given that both grade I and grade II hypothalamic damage involve the anterior hypothalamus, differences in oxytocin concentrations between patients with different grades of hypothalamic lesions was not anticipated; yet these findings were likely due to the impact of the small sample sizes (n=6 to 14 per group) on obtaining a reliable estimate of oxytocin concentrations for each grade of hypothalamic damage, and consequent lack of statistical power.

In studies assessing change in oxytocin concentrations pre- and post-intervention, significant differences between craniopharyngioma and controls were observed for studies implementing exercise stimulation, but not those using prandial intervention. Specifically, Daubenbüchel et al. (2016) did not find any significant differences when comparing the change between pre- and post-prandial salivary oxytocin concentrations between craniopharyngioma and controls, with both groups showing similar post-prandial (compared to pre-prandial) decreases in oxytocin concentrations, suggesting that the postprandial oxytocin response is intact in craniopharyngioma. This pattern of post-prandial decrease in salivary oxytocin concentrations is consistent with a study that reported a decrease in plasma oxytocin concentrations at 30 and 60 minutes following a standardised mixed meal in healthy females (independent of age, calorie intake, and menstrual phase) (Aulinas et al., 2019). By contrast, Gebert et al. (2018) found that in response to exercise all patients with craniopharyngioma showed a decrease in salivary oxytocin concentrations, compared to pre-exercise concentrations (−13.7%), whilst controls showed an increase (+24.8%) as expected. This was similarly found for the subsample of this study reported in Brandi et al. (2020), with a 7.90% decrease in oxytocin concentrations in patients with craniopharyngioma, compared to a 21.26% increase in controls.

Overall, findings from studies focusing on the endogenous oxytocin system suggest that deficits may only be identified when assessing the reactivity of the oxytocin system to stimulation (e.g., exercise), as opposed to single measurements of baseline concentrations alone. Therefore stimulation paradigms may pose an appropriate methodology for assessing the integrity of the oxytocin system in craniopharyngioma.

### 3.2 Associations between the oxytocin system and key outcomes

Current evidence suggests that dysregulated oxytocin in craniopharyngioma is associated with BMI (Daubenbüchel et al., 2016; Daubenbüchel et al., 2019; Gebert et al., 2018) and affective function (Gebert et al., 2018; Özyurt et al., 2020), while there are mixed findings for a relationship with social cognition (Brandi et al., 2020; Hoffmann et al., 2017; Özyurt et al., 2020).

#### 3.2.1 Oxytocin, metabolism, and eating behaviours

Convergent findings from two independent studies suggest a relationship between the change in pre- and post-stimulation concentration of salivary oxytocin and BMI and eating behaviours in craniopharyngioma. Specifically, Daubenbüchel et al. (2016) found that higher BMI was associated with smaller pre- vs. post-prandial decrease in salivary oxytocin concentrations in patients, whilst no association was found for controls. It is important to note that the authors did not report descriptive data on BMI, and therefore it is unknown how the dispersion of BMI values within the control group might have accounted for the lack of association here. It was additionally found that a smaller change in salivary oxytocin concentration was associated with subjective eating behaviours in patients with craniopharyngioma, specifically, with increased concerns about eating and weight (Daubenbüchel et al., 2019). Moreover, Gebert et al. (2018) found that across the whole sample, participants with higher BMI showed a smaller increase in salivary oxytocin concentrations post-exercise stimulation than those with lower BMI.

Given that higher BMI was associated with blunted changes in salivary oxytocin concentrations following prandial and exercise intervention, BMI may pose a key metabolic feature moderating oxytocin dysregulation in this group.

#### 3.2.2 Oxytocin and neurobehavioural impairment

A complex association was observed between salivary oxytocin and anxiety, with different relationships found for oxytocin with state anxiety (i.e., the transient response to a psychosocial stressor) and trait anxiety (i.e., the tendency to feel anxious across different contexts). Specifically, Gebert et al. (2018) found that greater trait anxiety was associated with higher baseline salivary oxytocin, whilst blunted release of oxytocin (i.e., no/ a smaller increase between pre- and post-exercise salivary oxytocin concentrations) was a significant predictor of greater state anxiety in craniopharyngioma. In addition, Özyurt et al. (2020) found that lower baseline salivary oxytocin concentrations were associated with greater state anxiety and severity of depression symptoms, across the whole sample. Stress-inducing contexts are known to increase the secretion of oxytocin (Takayanagi & Onaka, 2021), which can have an anxiolytic effect; when considering trait anxiety, it may be that over time, repeated initiation of oxytocin release due to heightened trait anxiety results in the downregulation of oxytocin receptors, and thus, an increase in circulating oxytocin concentrations (Uzun et al., 2022), possibly accounting for the positive relationship between baseline oxytocin and trait anxiety. Across the two studies however, lower concentrations of baseline oxytocin (Özyurt et al., 2020) and a blunted oxytocin release in response to exercise (Gebert et al., 2018) were associated with higher state anxiety. The association between oxytocin dysregulation and anxiety therefore likely poses a complex relationship that is dependent on the context and type of anxiety, and will benefit from advances in standardised tools and protocols, to facilitate meta-analysis and investigation of moderators related to the paradigm or conditions of the studies.

Limited evidence for an association between baseline oxytocin and socio-cognitive functioning emerged. Specifically, no associations between baseline salivary oxytocin concentrations and empathy quotient scores or socio-cognitive tasks, such as the Reading the Mind in the Eyes Test (Brandi et al., 2020), Theory of Mind (as measured by the Movie Assessment of Social Cognition), and Identification of Emotional Expressions in Voices (Özyurt et al., 2020) were observed, across the whole sample. Patients with hypothalamic damage, however, were found to have reduced Theory of Mind and reduced accuracy in identifying emotional vocal expressions compared to controls; yet, this difference was not observed when comparing all patients (i.e., no hypothalamic damage and hypothalamic damage) with controls (Özyurt et al., 2020). No group differences in baseline salivary oxytocin concentrations were found in this study (Özyurt et al., 2020). This suggests that hypothalamic damage may pose a mechanism underlying socio-cognitive difficulties, independent of effects of baseline oxytocin. The mechanisms underlying socio-cognitive difficulties in craniopharyngioma therefore remain to be established, and further research is required in order to delineate the specific direct and indirect effects of hypothalamic damage on this relationship. We should note here a methodological concern regarding the assessment of associations between oxytocin and neurobehavioural or metabolic outcomes by pooling participants across patient and control subgroups, in the presence of mean group differences in the associated variables (Gebert et al., 2018; Özyurt et al., 2020), as this practice may result in illusory correlations (Hassler & Thorsten, 2003). A more appropriate approach would be to pool correlation coefficients across samples (Hassler & Thorsten, 2003).

## 4 Discussion

This systematic review provides preliminary evidence that dysregulation of the oxytocin system may be associated with neurobehavioural functioning and BMI, and therefore, may pose a mechanism underlying these features in craniopharyngioma. While no significant differences were found between baseline salivary oxytocin concentrations in patients and controls, the findings of this review suggest that patients with craniopharyngioma may present a deficit in oxytocin secretion in response to a stressor, and that hypothalamic damage poses a likely moderator of the severity of this dysregulation. However, the methods of measuring endogenous oxytocin implemented by these studies (e.g., sampling type, quantification assay; see Tabak et al. (2023)) may not be sensitive to identifying differences in baseline concentrations between patients and controls, highlighting the need for the utilisation of more valid measurement protocols in future research.

A number of limitations in the methods implemented by the studies in the present review may account for the presence or absence of significant differences in baseline oxytocin concentrations between patients and controls. First, the studies used single salivary samples for oxytocin collection, yet the physiology (i.e., the diffusion into and clearance from saliva) of salivary oxytocin has not been established and its association with circulatory plasma and cerebrospinal fluid (CSF) oxytocin is unknown, and therefore may not present a valid trait marker of the central oxytocin system (Martins et al., 2020). Plasma has been identified as the favourable alternative to CSF measures, as the normal physiological range of < 10 pg/mL in mammal circulation has been established for extracted samples quantified using RIA (Leng & Sabatier, 2016). No studies in this review collected plasma oxytocin highlighting a key limitation of current research in craniopharyngioma in need of implementation. Second, one research group (Daubenbüchel et al., 2016) used EIA rather than RIA to quantify oxytocin concentrations, despite EIA having been criticised for having low sensitivity and high inter-assay variability (Tabak et al., 2023). The need for assays of higher sensitivity is especially relevant for patients with craniopharyngioma, since it is anticipated that this condition may have lower concentrations of central and/or peripheral oxytocin. Lastly, irrespective of the issues surrounding salivary sampling and quantification assays, single baseline measures of peripheral oxytocin concentrations have shown variability in the same individual at the same time, across different days, and therefore it is questionable whether single baseline samples can provide a valid and reliable marker of the integrity or function of the oxytocin system (Martins et al., 2020). The absence of studies collecting plasma oxytocin and the inconsistent implementation of RIA quantification methods, limits the validity of existing assessments of the endogenous oxytocin system. Additionally, given the intra-individual variability of oxytocin concentrations (Martins et al., 2020), alternative sampling protocols to single samples are necessary to reliably assess the endogenous oxytocin system in this population. At present, these limitations, combined with the lack of a standardised range of baseline values that could be utilised to assess oxytocin deficiency in routine care, emphasise a key area in need of investigation.

Repeated sampling and stimulation paradigms may prove more suitable methods for limiting the effects of intra-individual variability and increase the validity of baseline measurements (Martins et al., 2020) when assessing oxytocin concentrations in craniopharyngioma. For example, stimulation paradigms may present a more valid method of characterising the endogenous oxytocin system than single baseline measurements, with meta-analytic evidence supporting a relationship between peripheral and CSF measurements in response to stress stimulation in animals (i.e., pre- and post-stress stimulation measurements of oxytocin) (Valstad et al., 2017). In humans, stimulation by exercise, sexual self-stimulation, and psychosocial stress have been established to initiate a robust and reliable, fast-acting increase in salivary oxytocin concentrations (Alley et al., 2019; Jong et al., 2015) and plasma oxytocin concentrations (Carmichael et al., 1987; Hew-Butler et al., 2008; Pierrehumbert et al., 2010). Research work has additionally suggested 3,4-methylenedioxymethamphetamine (MDMA) administration as an effective provocation test for identifying a clinically meaningful oxytocin deficiency in patients with central diabetes insipidus (i.e., vasopressin deficiency) (Atila et al., 2023). In the present review, a difference in endogenous oxytocin concentrations between patients and controls was only found in one research group (Brandi et al., 2020; Gebert et al., 2018) assessing the reactivity of the oxytocin system in response to exercise stimulation. Therefore, stimulation paradigms provide evidence that the oxytocin system may be compromised in craniopharyngioma. Future research implementing stimulation paradigms (Valstad et al., 2017) and/ or repeated sampling of plasma and/or salivary oxytocin (at baseline) (Martins et al., 2020) are required in order to sensitively and accurately characterise the physiology of the oxytocin system in craniopharyngioma. In addition, oxytocin has been shown to elicit a diurnal rhythm with peak CSF concentrations at 12:00 (Amico et al., 1983), and more recently, to have a pulsatile architecture of secretion during resting state (Baskaran et al., 2017). Investigation into the dynamics of peripheral oxytocin secretion at rest, using repeated sampling across a single day/ night, may therefore identify deficits at specific phases of the cycle that cannot be captured using measurements at single time points, highlighting another area in need of exploration in patients with craniopharyngioma.

Differences in baseline oxytocin concentrations were identified between patients depending on presence of hypothalamic damage, suggesting hypothalamic damage as a probable moderator of oxytocin dysregulation in craniopharyngioma. Specifically, patients with hypothalamic damage were found to have lower salivary oxytocin concentrations than patients with no hypothalamic damage (Daubenbüchel et al., 2016; Gebert et al., 2018). Hypothalamic damage implicates the anterior hypothalamus, where oxytocin synthesising neurons of the paraventricular and supraoptic nuclei send axonal projections to the posterior pituitary for the release of oxytocin in peripheral circulation, as well as axonal projections and collaterals to central targets (Althammer & Grinevich, 2017). Therefore, patients with craniopharyngioma and hypothalamic damage may present either with an oxytocin-synthesising deficiency, and/or a deficit in oxytocin section if axonal projection routes are disrupted (Gebert et al., 2018). The effect of hypothalamic damage on the oxytocin system therefore needs to be disentangled before firm conclusions on the mechanisms of oxytocin dysregulation in craniopharyngioma may be established.

Hypothalamic damage was assessed by the studies in this review by visual inspection of the presence or absence of lesions to the anterior and/or posterior (with or without mamillary body involvement) hypothalamus (Müller et al., 2012). While the Müller grading system is clinically well-established, it does not allow for the assessment of parameters such as volume, or possible microstructure alterations that may impact the function of the oxytocin system. A volumetric approach to measuring hypothalamic damage in patients with craniopharyngioma has been explored, where lower hypothalamic volume was associated with greater fat mass and higher leptin levels (Fjalldal et al., 2019), suggesting hypothalamic volume as a quantitative marker of metabolic dysfunction in this condition. Existing methods may also be enhanced by implementing advanced imaging sequences, such as diffusion neurite orientation dispersion and density imaging (NODDI) and automated hypothalamic segmentation tools (Billot et al., 2020), that may be more sensitive to pathological change and allow for better characterisation of hypothalamic involvement. Utilisation of quantitative parameters to assess hypothalamic damage therefore presents an important avenue in need of exploration when investigating the oxytocin system and neurobehavioural and metabolic outcomes, in patients with craniopharyngioma.

While the extent to which the oxytocin system is compromised in craniopharyngioma is not yet clear, supplementation with exogenous oxytocin poses a potential therapeutic avenue, by restoring the inflammatory activation caused by the cystic and solid components of craniopharyngioma. In vivo evidence has shown that oxytocin pre-treatment reduced the inflammatory microenvironment generated by ox-Low Density Lipoprotein (ox-LDL) in the hippocampus of mice with craniopharyngioma, which subsequently improved cognitive function as measured by faster escape latencies in the Morris Water Maze Test (Wang et al., 2023). Anti-inflammatory effects have similarly been observed in obese mice where oxytocin infusion was found to reduce inflammation of visceral adipose tissue, reduce peripheral markers of systemic inflammation such as amyloid A levels, and increase circulating levels of adiponectin (an anti-inflammatory marker) (Szeto et al., 2020). The potential therapeutic benefits of the central and peripheral anti-inflammatory effects of exogenous oxytocin in craniopharyngioma need to be investigated in future research.

Studies addressing the direct effects of exogenous oxytocin on behavioural and/or metabolic outcomes in patients with craniopharyngioma are sparse, do not adhere to strict standards (e.g., randomization, double blinding, placebo-controlled) and mixed in methods (e.g., the length of treatment, dosage of intranasal oxytocin, and the outcomes measured) and participant characteristics, which impedes drawing any firm conclusions. Some promising evidence exists from a recent randomised placebo-controlled crossover pilot trial of intranasal oxytocin (16 to 24IU, three times per day) over eight weeks in 10 patients aged 10-35 years with hypothalamic obesity (secondary to a hypothalamo-pituitary tumour) (McCormack et al., 2023). No improvements in BMI were reported, but benefits for anxiety and impulsive traits were found (McCormack et al., 2023), suggesting exogenous oxytocin may indeed have benefits for neurobehavioural functioning in conditions affecting the hypothalamic region. Repeated administration (Terenzi & Ingram, 2005) and long-term use (Du et al., 2017) of oxytocin have been suggested to desensitise the oxytocin system (and consequently lead to worsened outcomes) (Du et al., 2017), suggesting that dose and administration frequency are important parameters. It has been shown that intermittent dosing (i.e., every other day) was more effective for attenuation of neural reactivity in subjects with elevated anxiety, compared to dosing every day (Kou et al., 2022). Studies with larger sample sizes, control conditions, double-blinding, and investigating optimal dosage schedules, are therefore required before implementation of exogenous oxytocin or analogues (e.g., Carbetocin; (Roof et al., 2023)) in routine care of craniopharyngioma can be supported.

### 4.1 Limitations

This systematic review aimed to assess the oxytocin system and its associations with neurobehaviour and metabolic parameters in craniopharyngioma. It should however be noted that this review could have been more inclusive of other tumours known to affect the hypothalamo-pituitary region such as prolactinoma and pilocytic astrocytoma, as included in the samples of two other related studies (Daughters et al., 2017; McCormack et al., 2023). To maximise homogeneity and control for the potential confound of additional clinical features associated with other tumours, only studies assessing patients with craniopharyngioma were included in the present review. Despite this restriction, high heterogeneity among the studies in relation to the sample demographics (e.g., age distribution) and methods of assessing the oxytocin system (e.g., sampling type, sampling protocol, quantification assay) was observed. As a result, it was not possible to statistically assess the relative contribution of potential confounding factors such as hypothalamic damage, age, and sex. Moreover, this review only included five independent samples, with the majority of studies reporting on the same, or a sub-group of the same, sample; our review is therefore based on the data of a limited number of independent observations. While the paucity of independent studies is likely contributed to by the rarity of craniopharyngioma (0.5 to 2 cases per million people per year (Nielsen et al., 2011), further clinical studies with larger sample sizes are needed.

Another important limitation is the broad age range of participants included in this review. A distinct pattern of *OXTR* expression across the lifespan has been reported with peak *OXTR* expression being identified during childhood; in particular, this increased expression was found in the mediodorsal nucleus of the thalamus, which has been associated with attention and memory (Pergola et al., 2018). *OXTR* expression then decreases during adulthood, until a second peak in late adulthood (Rokicki et al., 2022). Studies investigating the oxytocin system in craniopharyngioma may therefore need to restrict study samples to specific age groups in order to more sensitively assess any underlying relationships between oxytocin and socio-cognitive and behavioural outcomes.

## 5 Conclusions

Overall, this review suggests that patients with craniopharyngioma experience dysregulation of the oxytocin system. The presence of hypothalamic damage may constitute a key moderator of oxytocin dysregulation in craniopharyngioma and subsequent affective and metabolic related outcomes. It however remains challenging to draw firm conclusions from the current literature, particularly due to the lack of valid and reliable oxytocin sampling and quantification methods, the limited number of studies, and the heterogeneity of the study designs. Future research implementing appropriate assessments of peripheral oxytocin concentrations are required in order to understand the mechanisms underlying oxytocin dysregulation in craniopharyngioma (i.e., is there an oxytocin-synthesising deficiency and/or a release deficit). Once valid protocols for measurement of oxytocin have been established, multi-centre studies measuring oxytocin concentrations in craniopharyngioma and its relationship with neurobehavioural outcomes, eating behaviours, and metabolic outcomes may delineate which subgroups are at greater risk of presenting oxytocin dysregulation. Additionally, if craniopharyngioma is shown to be associated with oxytocin insufficiency, this finding will lay the foundations for future research into exogenous oxytocin as a therapeutic in this population, once optimal dosing and administration protocols have been established.

## Data Availability

All data produced in the present work are contained in the manuscript.

## Author Contributions

Amy Mann: Conceptualization; Methodology; Writing- original draft; Writing – reviewing and editing. Jennifer Kalitsi: Methodology. Khushali Jani: Methodology. Daniel Martins: Conceptualization; Writing- reviewing and editing. Ritika Kapoor: Conceptualization; Supervision; Writing- reviewing and editing. Yannis Paloyelis: Conceptualization; Supervision; Writing- reviewing and editing.

## Ethical Considerations

Not applicable.

